# Assessment of Retrospective Collection of EQ-5D-5L in US Patients with COVID-19

**DOI:** 10.1101/2023.01.18.23284602

**Authors:** Xiaowu Sun, Manuela Di Fusco, Laura Puzniak, Henriette Coetzer, Joann M. Zamparo, Ying P. Tabak, Joseph C. Cappelleri

**Author notes:** **Corresponding author:** Xiaowu Sun, Clinical Trial Services, CVS Health, Woonsocket, RI, USA.

## Abstract

**Background:** The impact of COVID-19 goes beyond the acute phase of infection. It is imperative to evaluate health related quality of life (HRQoL) pre-COVID-19, but there is currently no evidence of the retrospective application of the EQ-5D-5L for COVID-19 studies.

**Methods:** Subjects with ≥1 self-reported symptom and positive RT-PCR for SARS-CoV-2 at CVS Health US test sites were recruited between 01/31/2022-04/30/2022. On the day of enrollment which was around day 3 after testing positive, consented participants completed the EuroQol 5D-5L (EQ-5D-5L) questionnaire twice : a modified version where all the questions were past tense to retrospectively assess pre-COVID-19 baseline QoL, and the standard version in present tense to assess current HRQoL. Duncan’s new multiple range test was adopted for post analysis of variance pairwise comparisons of EQ-VAS means between problem levels for each of 5 domains. A linear mixed model was applied to check whether the relationship between EQ visual analog scale (VAS) and utility index (UI) was consistent pre-COVID-19 and during COVID-19. Matching-adjusted indirect comparison was used to compare pre-COVID-19 UI and VAS scores with those of the US population. Cohen’s d was used to quantify the magnitude of difference in means between two groups.

**Results:** Of 676 participants, 10.2% were age 65 or more years old, 73.2% female and 71.9% white. Diabetes was reported by 4.7% participants and hypertension by 11.2%. The pre-COVID-19 baseline mean UI was 0.924 and the mean VAS was 87.4. The estimated coefficient for the interaction of UI-by-retrospective collection indicator (0=standard prospective collection for Day 3 after COVID-19 testing, 1=retrospective for pre-COVID-19), -4.2 (SE: 3.2), P=0.197, indicates that retrospective collection does not significantly alter the relationship between EQ-VAS and UI. After adjusting for age, gender, diabetes, hypertension, and percent of mobility problems, predicted means of pre-COVID-19 baseline VAS and UI were 84.6 and 0.866, respectively. Both of these means were close to published US population norms (80.4 and 0.851) than those observed (87.4 and 0.924). After adjusting for age, gender, diabetes, and hypertension, 19.0% patients with COVID-19 had mobility problems, which was significantly lower than US population norm 25.2%, P<0.001. The calculated ES for UI and VAS were 0.15 and 0.39, respectively.

**Conclusion:** At a group level the retrospectively collected pre-COVID-19 EQ-5D-5L is adequate and makes it possible to directly evaluate the impact of COVID-19 on HRQoL. Future studies are encouraged that are tailored to directly compare standard prospective assessment with retrospective assessment on the EQ-5D-5L during pre-COVID-19.

## Introduction

The impact of COVID-19 extends beyond clinical outcomes. In order to understand holistically the burden of COVID-19, it is important to measure the impact on quality of life. The EuroQoL Group 5 dimension and 5 level (EQ-5D-5L) instrument [1] is an internationally validated questionnaire that is widely used for measuring health-related quality of life (HRQoL) and deriving utilities for estimation of Quality Adjusted Life Years (QALYs). Retrospective utilization of the EQ-5D-5L to assess pre-event status is limited, but the feasibility and validity of its retrospective collection (EQ-5D-5L) to assess past HRQoL was investigated in prior studies [2, 3]. In Lawson et al (2020) [2], EQ-5D-5L was collected prospectively in patients 2 weeks prior to their date of elective hip or knee arthroplasty surgery and then retrospectively collected following their operation to recall their pre-operative health status. At a group level the agreement was high between prospective and retrospective measurements indicating retrospective collection could be valid in orthopedic clinical context.

Another study by Rajan et al (2021) [3] utilized the EQ-5D-5L prospectively among patients with a stroke at 3 months post-discharge and compared those results with the retrospective collection of their 3rd month EQ-5D-5L at 6th, 9th, or 12th month after hospital discharge. Considerable agreement was observed in mobility, self-care, and usual activities dimensions of EQ-5D-5L based on weighted kappa (range 0.72-0.95), and concordance was good to excellent for utility score based on intraclass correlation coefficients (range 0.79-0.81). The authors concluded that retrospective collection of EQ-5D-5L could be a valid alternative for assessing morbid health status.

Retrospective and prospective collection of EQ-5D-5L have been used in infectious disease PRO studies to establish the baseline pre-infection health status and the morbid health status over the course of the infection [4, 5]. However, the validity of retrospective collection of EQ-5D-5L was not assessed in those studies. A recent study utilizing retrospective and prospective collection of EQ-5D-5L were used in a PRO study of US symptomatic outpatients with positive RT-PCR for SARS-CoV-2. EQ-5D-5L was used to establish, respectively, the baseline pre-COVID-19 health status and the health status at week 1 and week 4 following infection. Subsequently, the pre-infection and COVID-19 health statuses were compared to evaluate the impact of COVID-19 on HRQoL [6]. This subsequent methodology study utilizing the PRO data collected in the COVID-19 study tests the validity of the use of retrospective collection of EQ-5D-5L for establishing pre-infection status for the first time for COVID-19-related studies.

## Data and Methods

### Data source

In the COVID-19 PRO study [6], participants were recruited between 01/31/2022 and 04/30/2022 among US adult outpatients with ≥1 self-reported symptom and positive RT-PCR test for SARS-CoV-2 at CVS Health test sites (ClinicalTrials.gov NCT05160636). The analytic population for the Di Fusco et. al. study was limited to participants unvaccinated or receiving the BNT162b2 COVID-19 vaccine only. To assess the validity of the retrospective collection methodology, all patients that were enrolled were included [6].

### EQ-5D-5L

EQ-5D-5L addresses quality of life across five dimensions (mobility, self-care, usual activities, pain/discomfort, and anxiety/depression) and five levels (no problems, slight problems, moderate problems, severe problems, extreme problems/unable). These five domains were converted into the Utility Index (UI) using the US-based weights [7, 8]. Moreover, it addresses a general assessment of health, the EQ visual analog scale (VAS), via a 101-point Visual Analog Scale (VAS) ranging from 0=“Worst imaginable health state” to 100=“Best imaginable health state” [1, 7]. On the day of enrollment which was around day 3 after testing positive, consented participants completed the EQ-5D-5L questionnaire twice a modified version where all the questions were past tense to retrospectively assess pre-COVID-19 baseline HRQoL and, separately, the standard version in present tense to assess current HRQoL. These two versions were administered in random order to balance the potential responder bias due to the order of administration.

### Statistical methods

To support the validity of retrospectively collected EQ-5D-5L, here are the relevant assumptions. Firstly, lower VAS is associated with higher problem levels within each dimension of EQ-5D-5L. Secondly, pre-COVID-19 cohort can be considered as a sample from general population. After adjusting for major distributional difference, mean VAS should be close to population norm. Thirdly, retrospective collection does not modify the relationship between VAS and UI. And fourthly, retrospectively collected VAS is more reasonable than population norm when evaluating COVID-19’s impact on HRQoL.

Categorical variables were described by using frequency and percentages. Continuous variables were described by using means and standard deviations. The analysis of variance (ANOVA) was used to test differences in means between different problem levels within each domain of EQ-5D-5L. Duncan’s new multiple range test was adopted for pairwise comparisons post ANOVA [9, 10], which may evaluate the association between VAS and problem levels.

A linear mixed model was used to characterize the relationship between EQ-VAS and UI [11]. The response variable was EQ-VAS. The explanatory variables included UI, the indicator for retrospective assessment variable RETRO (1=retrospective collection for pre-COVID-19, 0=standard collection for day 3 after COVID-19 testing) and its interaction with UI. The coefficient of the interaction term reflects the magnitude of relationship altered by retrospective assessment between EQ-VAS and UI. Random intercepts were incorporated to account for the clusters of assessments within participants.

Matching-adjusted indirect comparison (MAIC) [12] was used to compare pre-COVID-19 UI and VAS scores with those of the US population. To do that, weights for individuals in our sample were estimated so that weighted percentage of selected patient characteristics matched those published. Then two-sample t-tests were used to test differences between weighted EQ-VAS or UI with those in the US population, as reported in Jiang et al. (2021) [13].

The effect size (ES), Cohen’s d, was calculated to assess the magnitude of difference in means between two groups [14, 15]. Specifically, when comparing COVID-19 to pre-COVID-19 baseline, the ES was calculated as mean score differences divided by the standard deviation of score difference. When comparing to COVID-19 to US norm, the ES was calculated as the difference in mean COVID-19 score and US norm, divided by the pooled standard deviation of scores. Values of 0.2, 0.5, and 0.8 standard deviation (SD) units represent small, medium, and large ES, respectively. These cut-off estimates have been widely used to establish important differences in HRQoL studies [16].

## Results

Of 676 participants, 10.2% were age 65 or more years old, 73.2% female and 71.9% white. Asthma or chronic lung disease, diabetes, and hypertension were reported by 8.6%, 4.7%, and 11.2% participants, respectively. (Table 1)

**Table 1.** Retrospective Collection of EQ-5D-5L Utility Index (US Preference Weights) and EQ-VAS by Respondent Characteristics

|  | Index Score |  | VAS |  |
| --- | --- | --- | --- | --- |
|  | n (%) | Mean (SD) | n (%) | Mean (SD) |
| All | 676 | 0.924 (0.117) | 674 | 87.4 (10.9) |
| Age |  |  |  |  |
| 18-34 | 134 (19.8) | 0.905 (0.114) | 134 (19.9) | 88.4 (9.6) |
| 35-64 | 317 (46.9) | 0.932 (0.109) | 317 (47.0) | 87.6 (10.3) |
| 65+ | 69 (10.2) | 0.936 (0.106) | 68 (10.1) | 88.8 (9.2) |
| Gender |  |  |  |  |
| Female | 495 (73.2) | 0.920 (0.113) | 495 (73.4) | 87.2 (11.1) |
| Male | 181 (26.8) | 0.935 (0.125) | 179 (26.6) | 87.9 (10.3) |
| Race/Ethnicity |  |  |  |  |
| White or Caucasian | 486 (71.9) | 0.921 (0.118) | 484 (71.8) | 87.1 (10.7) |
| Black or African American | 32 (4.7) | 0.918 (0.166) | 32 (4.8) | 82.7 (14.8) |
| Hispanic | 85 (12.6) | 0.930 (0.105) | 85 (12.6) | 88.4 (11.4) |
| Asian | 35 (5.2) | 0.969 (0.051) | 35 (5.2) | 89.9 (9.0) |
| Patient Refused | 16 (2.4) | 0.927 (0.096) | 16 (2.4) | 86.6 (10.2) |
| Other | 22 (3.3) | 0.899 (0.131) | 22 (3.3) | 91.7 (9.1) |
| Chronic conditions |  |  |  |  |
| Asthma or Chronic Lung Disease | 58 (8.6) | 0.873 (0.152) | 57 (8.5) | 81.7 (14.0) |
| Diabetes | 32 (4.7) | 0.887 (0.131) | 32 (4.8) | 82.8 (11.1) |
| Hypertension | 76 (11.2) | 0.887 (0.158) | 75 (11.1) | 83.1 (12.2) |
| Extreme obesity | 23 (3.4) | 0.782 (0.258) | 23 (3.4) | 77.0 (13.7) |
| Immunocompromised Conditions or Weakened Immune System <sup>a</sup> | 27 (4.0) | 0.889 (0.134) | 27 (4.0) | 78.6 (15.2) |

<sup>a</sup> Immunocompromised conditions includes compromised immune system (such as from immuno-compromising drugs, solid organ or blood stem cell transplant, HIV, or other conditions), conditions that result in a weakened immune system, including cancer treatment, and kidney failure or end stage renal disease
VAS = Visual Analogue Scale
EQ = EuroQol

As shown in Figure 1, worse VAS was associated with a higher level of severity in each EQ-5D-5L domain, for both retrospectively collected pre-COVID-19 baseline and standard collection for day 3 after COVID-19 positive testing. A detailed summary of EQ-VAS by EQ-5D-5L domain level for retrospective collection for pre-COVID-19 and standard collection for COVID-19, as well as the pairwise comparisons between levels of each domain, are presented in Supplemental Table 1. For pre-COVID-19 mobility, mean EQ-VAS scores were 88.5, 75.9, 57.7 and 50.0 for those reporting no problem, slight problem, moderate problem, and severe problem, respectively. No participants reported ‘unable’ for mobility. According to Duncan’s tests, mean EQ-VAS was found significantly different among no problem, slight problem, and moderate/severe problem. In addition, a summary of EQ-VAS by health states with a sample size greater than 10 is presented in Supplemental Table 2.

**Table 2.**
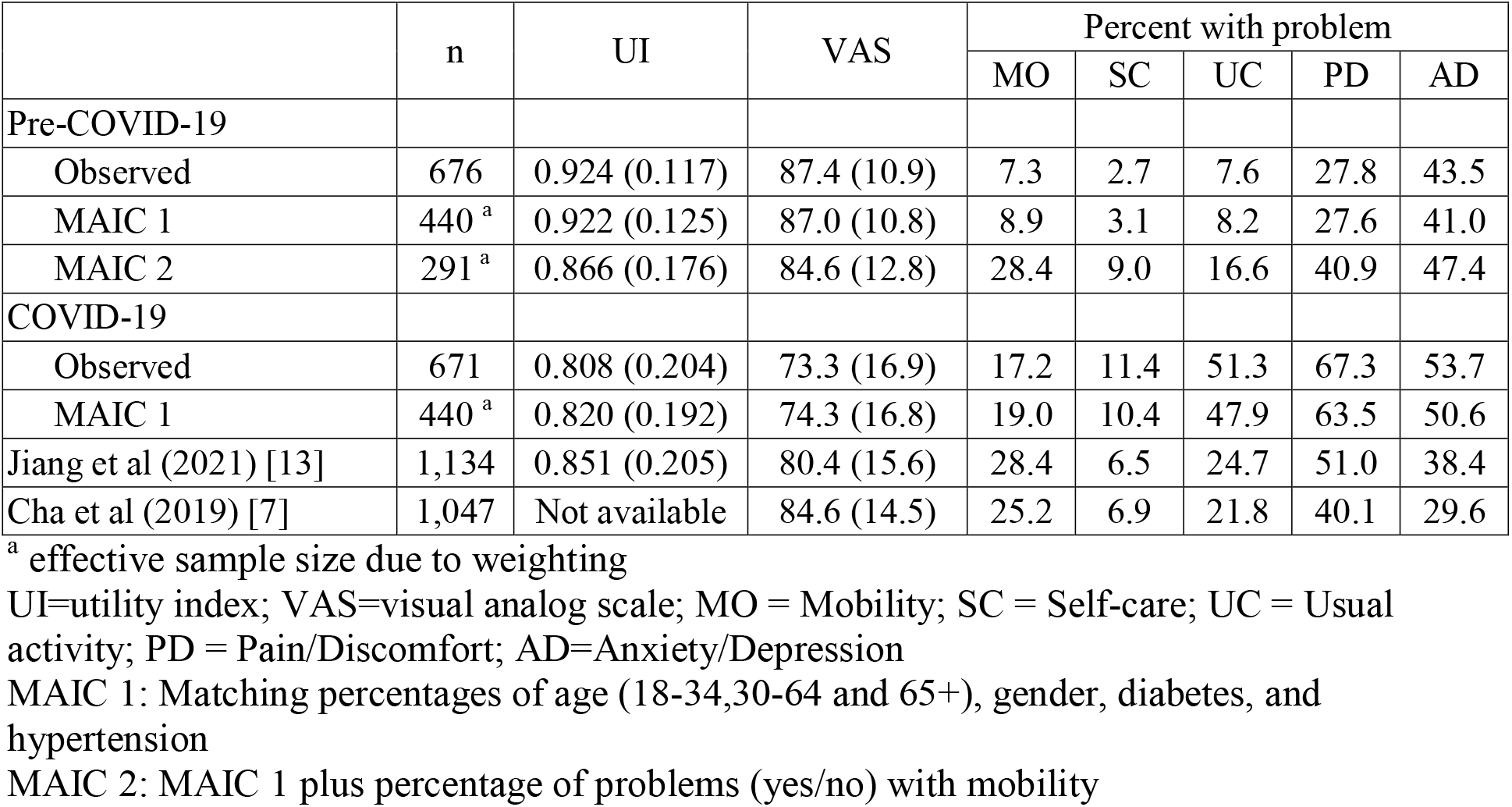
Observed and Weighted EQ-5D-5L Assessments with Comparisons with US Population Norms

|  | n | UI | VAS | Percent with problem |  |  |  |  |
| --- | --- | --- | --- | --- | --- | --- | --- | --- |
|  |  |  |  | MO | SC | UC | PD | AD |
| Pre-COVID-19 |  |  |  |  |  |  |  |  |
| Observed | 676 | 0.924 (0.117) | 87.4 (10.9) | 7.3 | 2.7 | 7.6 | 27.8 | 43.5 |
| MAIC 1 | 440 <sup>a</sup> | 0.922 (0.125) | 87.0 (10.8) | 8.9 | 3.1 | 8.2 | 27.6 | 41.0 |
| MAIC 2 | 291 <sup>a</sup> | 0.866 (0.176) | 84.6 (12.8) | 28.4 | 9.0 | 16.6 | 40.9 | 47.4 |
| COVID-19 |  |  |  |  |  |  |  |  |
| Observed | 671 | 0.808 (0.204) | 73.3 (16.9) | 17.2 | 11.4 | 51.3 | 67.3 | 53.7 |
| MAIC 1 | 440 <sup>a</sup> | 0.820 (0.192) | 74.3 (16.8) | 19.0 | 10.4 | 47.9 | 63.5 | 50.6 |
| Jiang et al (2021) [13] | 1,134 | 0.851 (0.205) | 80.4 (15.6) | 28.4 | 6.5 | 24.7 | 51.0 | 38.4 |
| Cha et al (2019) [7] | 1,047 | Not available | 84.6 (14.5) | 25.2 | 6.9 | 21.8 | 40.1 | 29.6 |

<sup>a</sup> effective sample size due to weighting
UI=utility index; VAS=visual analog scale; MO = Mobility; SC = Self-care; UC = Usual activity; PD = Pain/Discomfort; AD=Anxiety/Depression
MAIC 1: Matching percentages of age (18-34,30-64 and 65+), gender, diabetes, and hypertension
MAIC 2: MAIC 1 plus percentage of problems (yes/no) with mobility

**Figure 1.**
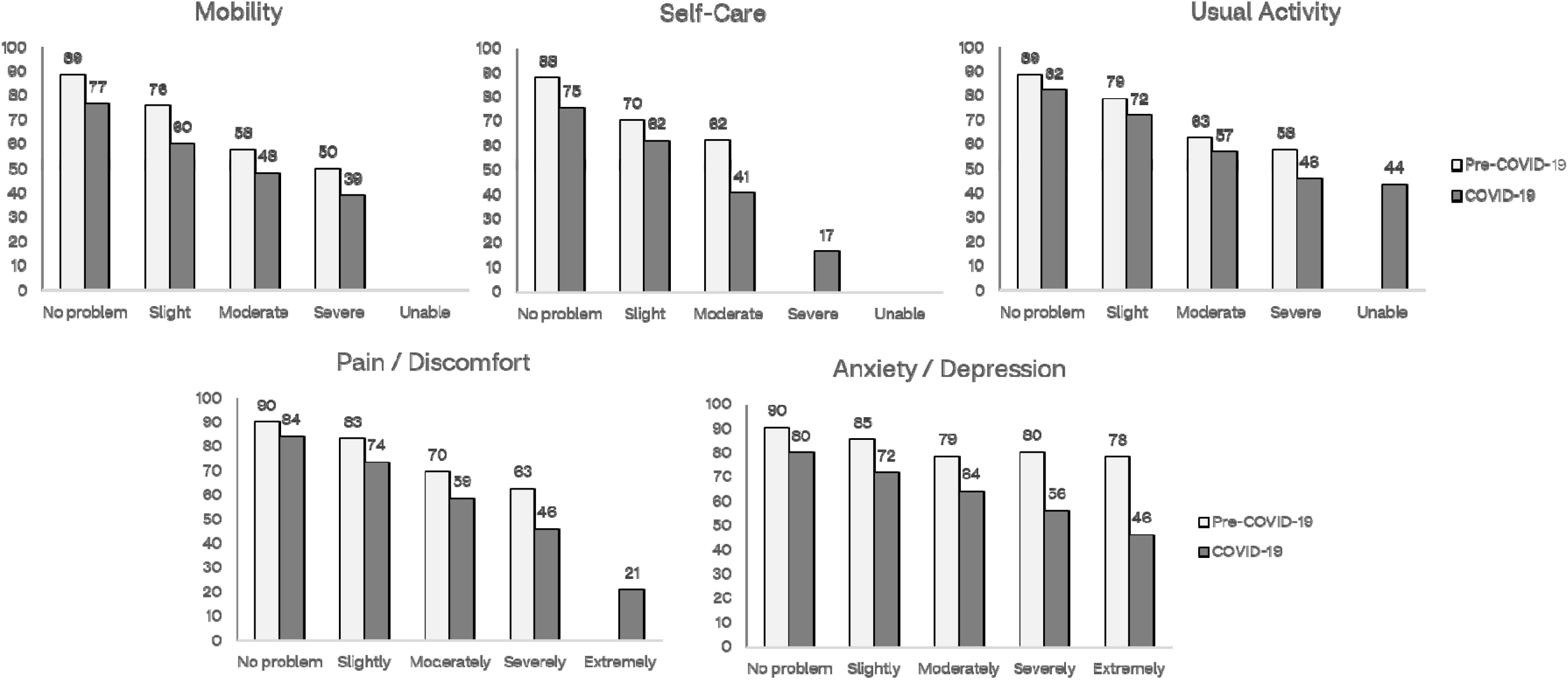
Sample Means of EQ-VAS by EQ-5D-5L Domain Level: Pre-COVID-19 and COVID-19

Compared with the US general population [13], the current cohort was predominantly female, white, and reported less chronic conditions. The pre-COVID-19 baseline mean utility index (UI) of 0.924 and mean VAS of 87.4 were higher than those in the US population (0.851 and 80.4, respectively). In addition, for all five domains, less problems were reported, which were prevalent in 7.1%, 6.7%, 7.9%, 27.3% and 43.5% of the current study cohort versus 28.4%, 6.5%, 24.7%, 51.0%, and 38.4% in Jiang et al (2021) [13] for mobility, self-care, usual activity, pain/discomfort and anxiety/depression, respectively.

For the model predicting EQ-VAS by using UI, RETRO and their interaction, the estimated coefficient of UI-by-RETRO interaction term was -4.2 (SE: 3.2), P=0.197, which indicated that the magnitude of relationship between EQ-VAS and UI was not significantly altered by retrospective assessment.

To compare with the US population norms for the EQ-5D-5L [13], we matched percentages of age category (18-29, 30-64 and ≥65 years), gender, diabetes, and hypertension in our sample to US population norms by the using MAIC approach [12]. After weighting, the effective sample size reduced to 440. The weighted mean of UI and VAS for pre-COVID-19 baseline decreased slightly to 87.0 and 0.922, respectively. Both were significantly higher than US population norms, P<0.001. When percent of problems with mobility was added, the effective sample size reduced to 291. The weighted mean of UI for pre-COVID-19 baseline became 0.866 (SD: 0.176), not statistically different from the US population norm 0.851 (P=0.253). The weight mean of VAS was 84.6 (SD:12.8) and higher than the US population norm of 80.4 (P<0.001). (Table 2)

Around day 3 after COVID-19 positive testing, UI and VAS were 0.808 and 73.3, respectively. When comparing with baseline assessment, Cohen’s d for UI and VAS were 0.68 and 1.01, a medium-to-large impact on UI and a large impact on VAS, respectively. When comparing with US population norms [13], Cohen’s d for UI and VAS were 0.21 and 0.44, a small impact on UI and a small-to-medium impact on VAS, respectively. However, the percent of problems with mobility, 17.2%, of COVID-19 cohort was significantly lower than 25.2% of US population [13], P<0.001. After matching on percentages of age category (18-29, 30-64 and ≥65 years), gender, diabetes, and hypertension, 19.0% with mobility problems was still significantly lower than 25.2%, P<0.001. The calculated ES for UI and VAS became 0.15 and 0.39, respectively. (Table 2)

## Discussion

In this current study implementing retrospective collection of EQ-5D-5L data, EQ-VAS generally declined along with more severe levels in each of 5 domains as expected. Strong empirical evidence indicates that retrospective collection did not materially alter the relationship between EQ-VAS and UI. After adjusting for demographic variables, chronic conditions, and percent of mobility problems, mean scores of EQ-VAS and UI approached those of US population norms. As with related studies which demonstrate the validity of retrospective collection of EQ-5D-5L through direct comparisons with standard collection [2, 3], results from the current study provide evidence to support the validity of retrospective EQ-5D-5L collection when it is not feasible to collect this information prospectively. To our knowledge, however, this is the first study of its kind in an adult outpatient COVID-19 cohort.

The pre-COVID-19 adjusted mean EQ-VAS of 84.6 is the same as that reported for the US general population in Cha et al (2019) [7], but still higher than that reported by Jiang et al (2021) [13]. Such difference may be due to underadjustment for participants’ characteristics and possible bias in retrospective assessment.

While useful to adjust for differences in participants’ characteristics between study cohort and the cohort deriving US population norms, there are limitations with MAIC. In the current study chronic conditions were reported less than US general population and Jiang et al (2021) [13].

Nevertheless, conditions cannot be adjusted that are either not commonly available in both studies or not consistently collected, which resulted in adjustment for diabetes and hypertension only. The addition of retrospectively collected problems with mobility for pre-COVID-19 in MAIC brought EQ-VAS and UI much closer to US population norm.

When it is not feasible to collect prospective measurements on EQ-5D-5L, as well as EQ-VAS and EQ-UI, the current evidence suggests that it reasonable to consider retrospective assessment of them under the situation found in our study, including the short recall period (only a few days) for the retrospective assessment. Especially for mobility, there is a high agreement between retrospective collection and standard collection [2, 3]. High percentage of no mobility problem is in alignment with the cohort comprising of people walking in the store for COVID-19 testing. Even so, we did not attempt to use all problem levels either too few to be reliably adjusted or unable to adjust for the level not observed. (Supplemental Table 1)

The retrospective assessment of EQ-VAS for pre-COVID-19 may either be recalled directly, or more likely be reconstructed based on current state of COVID-19 and the assumptions about probably change from pre-COVID-19 to COVID-19. The recall bias occurs because of errors and distortions in the recollection of pre-COVID-19 state as well as in the inferential process from pre-COVID-19 to COVID-19 [17]. As shown in Rajan et al (2021) [3], patients with a stroke tended to underestimate EQ-VAS retrospectively, which supports the ‘present state effect’ [17, 18], that is, a person who feels well might think his status improved and therefore tends to underestimate his previous state. Conversely, a patient with COVID-19 who feels bad might think that her status worsened and therefore tends to overestimate her pre-COVID-19 status.

Had EQ-5D-5L not been collected retrospectively, US population norms must be compared to evaluate COVID-19’s impact on HRQoL. The assumption is that COVID-19 causes more problems in mobility, self-care, usual activity, pain/discomfort and anxiety/depression, as well as overall health. However, current cohort of patients with COVID-19 had less problem with mobility than general population, whether matching on selected patient’s characteristics or not.

On the other hand, retrospectively collected baseline percentage of problems with anxiety and depression, 43.5%, was higher than US population norm, 38.4%, based on the cohort pre-pandemic [13], which is consistent with World Health Organization report about elevated prevalence of anxiety and depression during COVID-19 pandemic [19]. Comparisons of UI and VAS of patients with COVID-19 with population norms cannot account for such differences, which leads to unreliable estimates of COVID-19’s impact.

## Conclusion

At a group level the retrospectively collected pre-COVID-19 EQ-5D-5L is adequate when compared with US population norm and reasonably aligned when compared with standard collection of EQ-5D-5L for COVID-19. Retrospective collection of pre-COVID-19 EQ-5D-5L makes it possible to directly evaluate the impact of COVID-19 on health-related quality of life. Future studies are encouraged that are tailored to directly compare standard prospective assessment with retrospective assessment on the EQ-5D-5L during pre-COVID-19.

## Data Availability

Aggregated data that support the findings of this study are available upon reasonable request from the corresponding author XS, subject to review. These data are not publicly available due to them containing information that could compromise research participant privacy/consent.

## Declarations

### Ethics approval and consent to participate

This study was approved by the Sterling IRB, Protocol #C4591034. Participation in the study was voluntary and anonymous. Consent was obtained electronically via the CVS Health E-Consent platform. Participants were informed of their right to refuse or withdraw from the study at any time. Participants were compensated for their time.

### Consent for publication

All authors have given their approval for this manuscript version to be published.

## Authors’ contributions

All named authors meet the International Committee of Medical Journal Editors (ICMJE) criteria for authorship for this article. XS: Conceptualization, Methodology, Data curation, Formal analysis, Writing - original draft. MDF and YP: Investigation, Project administration, Conceptualization, Writing - Review & editing. LP, JMZ and HC: Conceptualization, Writing - Review & editing. JCC: Conceptualization, Methodology, Formal analysis, Writing - Review & editing.

## Acknowledgements

The authors acknowledge Alejandro Cane, Deepa Malhotra and Nancy Gifford (Pfizer employees), Joseph Ferenchick, Shiyu Lin and Shawn Edmonds (CVS Health employees) for specific contributions to this research project. Editorial support was provided by Laura Anatale-Tardiff and Leena Samuel at CVS Health and was funded by Pfizer.

---

**Supplemental Table 1.** Summary of EQ-VAS by EQ-5D-5L Domain Level: Retrospective Collection for Pre-COVID-19 and Standard Collection for COVID-19

|  | Pre-COVID-19 |  |  | COVID-19 |  |  |
| --- | --- | --- | --- | --- | --- | --- |
|  | n (%) | Mean (SD) | Duncan Grouping<sup>a</sup> | n (%) | Mean (SD) | Duncan Grouping<sup>a</sup> |
| All | 674 | 87.4 (10.9) |  | 671 | 73.3 (16.9) |  |
| Mobility |  |  |  |  |  |  |
| No problem | 626 (92.9) | 88.5 (9.5) | A | 558 (83.2) | 76.6 (14.3) | A |
| Slight | 40 (5.9) | 75.9 (13.6) | B | 88 (13.1) | 60.0 (19.3) | B |
| Moderate | 6 (0.9) | 57.7 (15.8) | C | 21 (3.1) | 48.0 (15.4) | C |
| Severe | 2 (0.3) | 50.0 (28.3) | C | 4 (0.6) | 39.0 (20.7) | C |
| Unable | 0 |  |  | 0 |  |  |
| Self Care |  |  |  |  |  |  |
| No problem | 656 (97.3) | 87.8 (10.3) | A | 596 (88.8) | 75.4 (15.3) | A |
| Slight | 17 (2.5) | 70.4 (16.8) | B | 60 (8.9) | 61.8 (17.0) | A |
| Moderate | 1 (0.1) | 62.0 (.) | B | 13 (1.9) | 40.6 (16.4) | B |
| Severe | 0 |  |  | 2 (0.3) | 16.5 (16.3) | C |
| Unable | 0 |  |  | 0 |  |  |
| Usual Activity |  |  |  |  |  |  |
| No problem | 621 (92.1) | 88.6 (9.6) | A | 329 (49.0) | 82.3 (11.9) | A |
| Slight | 36 (5.3) | 78.8 (12.8) | A | 207 (30.8) | 71.7 (13.0) | B |
| Moderate | 15 (2.2) | 62.5 (14.4) | B | 102 (15.2) | 56.7 (14.5) | C |
| Severe | 2 (0.3) | 57.5 (10.6) | B | 24 (3.6) | 45.9 (12.6) | D |
| Unable | 0 |  |  | 9 (1.3) | 43.7 (23.3) | D |
| Pain / Discomfort |  |  |  |  |  |  |
| No problem | 487 (72.3) | 90.1 (8.5) | A | 221 (32.9) | 84.2 (11.4) | A |
| Slightly | 146 (21.7) | 83.3 (10.5) | A | 307 (45.8) | 73.6 (12.9) | B |
| Moderately | 37 (5.5) | 69.7 (14.9) | B | 120 (17.9) | 58.6 (15.6) | C |
| Severely | 4 (0.6) | 62.8 (19.6) | B | 20 (3.0) | 46.0 (19.4) | D |
| Extremely | 0 |  |  | 3 (0.4) | 21.0 (13.9) | E |
| Anxiety / Depression |  |  |  |  |  |  |
| No problem | 381 (56.5) | 90.4 (9.0) | A | 312 (46.5) | 80.1 (13.6) | A |
| Slightly | 201 (29.8) | 85.5 (10.8) | A B | 204 (30.4) | 71.8 (15.3) | B |
| Moderately | 73 (10.8) | 78.6 (13.3) | B | 113 (16.8) | 64.3 (16.6) | B |
| Severely | 16 (2.4) | 80.3 (11.5) | B | 35 (5.2) | 56.0 (18.6) | C |
| Extremely | 3 (0.4) | 78.3 (8.3) | B | 7 (1.0) | 46.3 (28.9) | D |

<sup>a</sup> Means of EQ-VAS are significantly different between levels with different letters (P<0.05) according to Duncan's new multiple range tests|

**Supplemental Table 2.** Summary of EQ-VAS by Health States with Volume >10: Retrospective Collection for Pre-COVID-19 and Standard Collection for COVID-19

| Pre-COVID-19 |  |  | COVID-19 |  |  |  |
| --- | --- | --- | --- | --- | --- | --- |
| Health State | n | EQ-VAS Mean (SD) | Health State | n | EQ-VAS Mean (SD) | ARI Symptoms, Mean (SD) |
| 11111 | 316 | 91.8 (7.6) | 11111 | 130 | 87.8 (9.8) | 4.0 (2.0) |
| 11112 | 124 | 88.2 (9.3) | 11121 | 61 | 80.5 (9.9) | 4.7 (2.3) |
| 11122 | 45 | 84.5 (9.5) | 11221 | 56 | 74.3 (11.5) | 5.6 (2.2) |
| 11121 | 35 | 86.5 (11.4) | 11122 | 43 | 79.1 (9.8) | 5.3 (2.4) |
| 11113 | 30 | 84.3 (7.6) | 11112 | 37 | 83.7 (10.3) | 4.1 (2.4) |
| 11123 | 20 | 82.5 (7.7) | 11222 | 35 | 73.0 (9.8) | 5.3 (2.1) |
|  |  |  | 11223 | 21 | 72.7 (10.2) | 5.3 (2.7) |
|  |  |  | 11211 | 16 | 79.7 (9.5) | 5.0 (2.8) |
|  |  |  | 11113 | 12 | 77.9 (8.1) | 4.6 (3.0) |
|  |  |  | 11123 | 11 | 71.8 (15.0) | 4.8 (2.0) |
|  |  |  | 11212 | 11 | 72.3 (18.5) | 4.2 (2.5) |

## References

[1] EuroQol Research Foundation. EQ-5D-5L User Guide, Version 3.0. https://euroqol.org/publications/user-guides Accessed August 5, 2021.

[2] Lawson A, Tan AC, Naylor J, Harris IA. Is retrospective assessment of health-related quality of life valid? BMC Musculoskeletal Disorders. 2020;21:1–10.

[3] Rajan SS, Wang M, Singh N, Jacob AP, Parker SA, Czap AL, et al. Retrospectively Collected EQ-5D-5L Data as Valid Proxies for Imputing Missing Information in Longitudinal Studies. Value Health. 2021;24:1720–7.

[4] Barbut F, Galperine T, Vanhems P, Le Monnier A, Durand-Gasselin B, Canis F, et al. Quality of life and utility decrement associated with Clostridium difficile infection in a French hospital setting. Health and Quality of Life Outcomes. 2019;17:1–7.

[5] Glick HA, Miyazaki T, Hirano K, Gonzalez E, Jodar L, Gessner BD, et al. One-Year Quality of Life Post-Pneumonia Diagnosis in Japanese Adults. Clin Infect Dis. 2021;73:283–90.

[6] Di Fusco M, Sun X, Moran MM, Coetzer H, Zamparo JM, Puzniak L, et al. Impact of COVID-19 and effects of BNT162b2 on patient-reported outcomes: quality of life, symptoms, and work productivity among US adult outpatients. Journal of Patient-Reported Outcomes. 2022;6.

[7] Cha AS, Law EH, Shaw JW, Pickard AS. A comparison of self-rated health using EQ-5D VAS in the United States in 2002 and 2017. Quality of Life Research. 2019;28:3065–9.

[8] Pickard AS, Law EH, Jiang R, Pullenayegum E, Shaw JW, Xie F, et al. United States valuation of EQ-5D-5L health states using an international protocol. Value in Health. 2019;22:931–41.

[9] Rosner B. Fundamentals of biostatistics. Eighth ed. Boston, MA: Cengage learning; 2015.

[10] Duncan DB. T tests and intervals for comparisons suggested by the data. Biometrics. 1975:339–59.

[11] Brown H, Prescott R. Applied mixed models in medicine: John Wiley & Sons; 2015.

[12] Signorovitch JE, Sikirica V, Erder MH, Xie J, Lu M, Hodgkins PS, et al. Matching-adjusted indirect comparisons: a new tool for timely comparative effectiveness research. Value in Health. 2012;15:940–7.

[13] Jiang R, Janssen MB, Pickard AS. US population norms for the EQ-5D-5L and comparison of norms from face-to-face and online samples. Quality of Life Research. 2021;30:803–16.

[14] Cohen J. Statistical power analysis for the behavioral sciences. 2nd ed. Hillsdale, NJ.: Lawrence Erlbaum Assoc; 1988.

[15] McLeod LD, Cappelleri JC, Hays RD. Best (but oft-forgotten) practices: expressing and interpreting associations and effect sizes in clinical outcome assessments. The American journal of clinical nutrition. 2016;103:685–93. Erratum: 2017; 105:241.

[16] Mouelhi Y, Jouve E, Castelli C, Gentile S. How is the minimal clinically important difference established in health-related quality of life instruments? Review of anchors and methods. Health and Quality of Life Outcomes. 2020;18:1–17.

[17] Blome C, Augustin M. Measuring change in quality of life: bias in prospective and retrospective evaluation. Value in Health. 2015;18:110–5.

[18] Kamper SJ, Ostelo RW, Knol DL, Maher CG, de Vet HC, Hancock MJ. Global Perceived Effect scales provided reliable assessments of health transition in people with musculoskeletal disorders, but ratings are strongly influenced by current status. Journal of clinical epidemiology. 2010;63:760-6. e1.

[19] World Health Organization. Mental Health and COVID-19: Early evidence of the pandemic’s impact. https://www.who.int/publications/i/item/WHO-2019-nCoV-Sci_Brief-Mental_health-2022.1 Accessed November 16, 2022.

